# Histopathology of extracutaneous tissues in a uremic calciphylaxis patient treated with human amnion-derived mesenchymal stem cells: a comparison with ESKD patients

**DOI:** 10.1101/2024.10.21.24315424

**Authors:** Cui Li, Shijiu Lu, Guang Yang, Yue Cao, Ming Zeng, Kang Liu, Yanggang Yuan, Yan Ding, Zhonglan Su, Fangyan Xu, Wenkai Ren, Wei Liu, Yi Xu, Jing Zhang, Xiaoxue Ye, Chunyan Jiang, Yugui Cui, Xiang Ma, Song Ning, Yujie Xiao, Changxing Luan, Qiang Ji, Zhiwei Zhang, Mufeng Gu, Changying Xing, Xiuqin Wang, Ningxia Liang, Feng Chen, Jiayin Liu, Lianju Qin, Youjia Yu, Ningning Wang

## Abstract

**Background:** Calciphylaxis, which mostly affects individuals with end-stage kidney disease (ESKD), is also known as calcific uremic arteriolopathy (CUA). It is a rare and fatal disease that manifests with calcification and thrombosis of microvessels, ischemia, and necrosis in skin tissues(ORPHA:280062). Histopathological features of extracutaneous tissues of CUA patients undergoing human amnion–derived mesenchymal stem cell (hAMSC) treatment remain unknown.

**Methods:** A female CUA patient, treated with hAMSCs for 20 months, passed away due to stroke. Histopathological features of her extracutaneous tissues were compared with those of ESKD patients (n = 7). Raman spectroscopy was applied to identify the composition of calcifications. The distribution of hAMSCs, derived from the amnion of a male fetus, in tissues of the CUA patient was determined by detecting the Y chromosome using reverse-transcription–polymerase chain reaction.

**Results:** Microvessel lesions were more prevalent in the extracutaneous tissues of the CUA patient than in those of ESKD patients, although the regenerated skin showed normal histological characteristics. The CUA patient exhibited calcifications of microvessel media, including the microvessels in the lungs, kidneys, spleen, pancreas, and uterus. Her mitral valve and kidney displayed severe calcification, identified as calcium phosphate with some calcium carbonate. hAMSCs were not detected in the tissues of the CUA patient.

**Conclusion:** Under the treatment strategy with hAMSCs, based on the effects of skin regeneration, microvascular lesions in the extracutaneous tissues of the CUA patient were more severe than those in ESKD patients. CUA should be considered a systemic disease when identifying treatment targets.

## Introduction

Calciphylaxis is a rare disease characterized by tissue ischemia and necrosis due to microvessel calcification, extravascular calcification, intimal fibroplasia, and thrombosis in the subcutaneous adipose tissues and dermis (ORPHA:280062). The patients have severe, painful cutaneous lesions with a median survival time of 3 months and a 1-year mortality rate of 80% due to infection^[1–4]^. Calciphylaxis is also known as calcific uremic arteriolopathy (CUA), given that most cases have been described in patients with end-stage kidney disease (ESKD)^[4–7]^. The presence of calcifications in small vessels has been reported in the extracutaneous tissues of calciphylaxis patients, which affects the prognosis^[4,8–15]^. However, it remains unknown whether calciphylaxis is a systemic process or it is confined to the skin tissues. Insights into the pathophysiology and identification of potential therapies have been limited due to the rarity of CUA^[16]^.

We have innovatively rescued a severe CUA patient using intravenous and local treatment with human amnion–derived mesenchymal stem cells (hAMSCs). The effects of hAMSC therapy included the inhibition of microvascular calcification, stimulation of angiogenesis, anti-inflammatory action, immune modulation, improvement of hypercoagulability, and restoration of integrity^[17,18]^. The tumorigenicity of cell therapy derived from induced pluripotent stem cells (iPSCs) for the treatment of diabetes has been documented^[19]^. However, the safety and effectiveness of hAMSC treatment of extracutaneous lesions in CUA patients remain uncertain.

With a more comprehensive and systematic assessment relative to antemortem findings, we compared the histopathological features of the extracutaneous tissues of a CUA patient treated with hAMSCs with those of ESKD patients. The calcium composition and distribution of hAMSCs in multiple tissues of the CUA patient were determined. Our findings provide a deeper understanding of the pathogenesis of extracutaneous lesions in calciphylaxis patients and further help improve the hAMSC therapy strategies for this devastating disease.

## Methods

### Study population

#### The female CUA patient underwent hAMSC therapy

A 30s female patient had been on peritoneal dialysis (PD) for 5 years and switched to hemodialysis (HD) due to PD-related peritonitis and decreased dialysis adequacy, with a weekly KT/V urea of 1.404. The CUA patient was admitted to our hospital in July 2018, because of progressive skin lesions with induration, plaques, purpura, reticulate livedo, and ecchymosis on her back, thighs, lower limbs, and buttocks. The patient was diagnosed with CUA, as described in our previous studies^[17,18]^.

Treatment with hAMSCs was approved because of worsening clinical manifestations and unresponsiveness to conventional calciphylaxis therapies, including the intravenous injection of sodium thiosulfate^[17,18]^. This study was approved by the Ethics Committee of the First Affiliated Hospital with Nanjing Medical University in China (2018-QT-001).

hAMSCs derived from human male fetuses (XY) were administered intravenously at a dosage of 1.0 × 10^6^ cells/kg in September 2018. They were also combined with local intramuscular injection along the wound edge (2.0 × 10^4^ cells/cm^2^) and external application of hAMSC-conditioned medium on the wound surface for a total of 15 months. Compared with pre-therapy, multiple skin lesions were successfully healed after 1 year, including those on the back, thighs, lower limbs, and buttocks^[18]^. Unfortunately, the patient died of a cerebral hemorrhage following cerebral infarction after hAMSC treatment for 20 months. Her parents donated her body for medical research and provided authorization for the research. We conducted an autopsy in compliance with Chinese laws.

#### The ESKD patients without calciphylaxis

To identify the pathological characteristics of the extracutaneous tissues of the CUA patient treated with hAMSCs, the data of the ESKD patients were collected from the archives of the Department of Forensic Medicine, Nanjing Medical University between January 1, 2007, and July 30, 2023. Informed consents were obtained from the immediate relatives of the patients.

#### Inclusion criteria

The inclusion criteria of the patients were as follows: (1) age between 18 and 70 years; (2) the diagnostic criteria for ESKD were met, based on the “KDIGO 2017 Clinical Practice Guideline Update for the Diagnosis, Evaluation, Prevention, and Treatment of Chronic Kidney Disease-Mineral and Bone Disorder (CKD-MBD)”^[20]^.

#### Exclusion criteria

ESKD patients with a history of calciphylaxis, allergic disease, serious infections, a history of malignancy, or poor-quality pathology specimens were excluded.

## Main observation indicators

### Clinical data from the CUA and ESKD patients

For the ESKD patients with and without calciphylaxis, basic information, such as age, gender, medical history, dialysis mode, cause of death, physical examinations, and laboratory examinations, was collected. For the CUA patient, imaging examinations, including ultrasound examination, computed tomography (CT) examination, and whole-body scan with ^99m^Tc-methylene diphosphonate (^99m^Tc-MDP), were recorded.

### Histopathological analysis

Extracutaneous tissues were obtained from ESKD patients (n = 7) and the CUA patient treated with hAMSCs. In the CUA patient, slides from extracutaneous tissues were stained with hematoxylin and eosin (H&E). In cases where calcification was identified through H&E staining, Alizarin red staining was used^[21]^. Furthermore, the presence of bacterial or fungal infections on the mitral valve was excluded through the application of H&E staining, periodic acid-Schiff staining^[22]^, and Grocott’s methenamine silver (GMS) staining^[23,24]^. Images were acquired using a light microscope (Olympus, BX53, Japan).

The pathological slides were evaluated by two experienced pathologists who were blind to the clinical information.

Parameters investigated in both groups encompassed microvessel calcification, intimal hyperplasia, fibrosis, interstitial calcium deposition, microthrombi, tumors, and abnormal tissue hyperplasia.

### Raman spectra and imaging

Formalin-fixed paraffin-embedded (FFPE) specimens from the mitral valve and kidneys of the CUA patient were analyzed using Raman spectroscopy as described previously^[25]^. The analysis was conducted using an inverted microscope with an external three-channel optical system (TI-U, Nikon, Japan), equipped with a 60× plane objective lens. The mitral valve and kidney tissue samples were placed on Raman Substrate Materials-CaF_2_ disks, and the CaF_2_ disks were mounted on the XY stage. The area of interest was selected using the brightfield image, and the instrument control software was used to automatically collect the Raman spectra of the tissues. To avoid high fluorescence, we chose an excitation wavelength of 785 nm, a laser power of 30 mW, and a spectrum acquisition time of 3 s to achieve a sufficient signal-to-noise ratio without causing any obvious tissue damage. Fast Raman imaging was performed with a 1000 nm step size and 3 s integration time per pixel. The Raman data analysis was performed using Labspec 6 software, and the baseline was subtracted based on polynomial fitting to produce a flat background.

### Detection of the human Y chromosome using RT-PCR

The DNA of the CUA patient was extracted from multiple extracutaneous tissues, including the brain, pituitary gland, parathyroid gland, spleen, lungs, cardiac valve, coronary artery, ovary, adipose tissue, skin of the thigh, and dermal tissue of the buttocks. The distribution of transplanted hAMSCs, derived from the amnion of a male fetus, in multiple tissues of the female CUA patient was assessed through the detection of the human Y chromosome using RT-PCR. The amplified primer sequences were derived from the sex-determining region Y (SRY primer sequences: SRY-609F, CCCGAATTCGACAATGCAATCATATGCTTCTGC; SRY-609R, CTGTAGCGGTCCCGTTGCTGCGGTG). Two healthy men and two healthy women were included as positive and negative controls, respectively. Figure 1 shows the study flow diagram.

**Figure 1:**
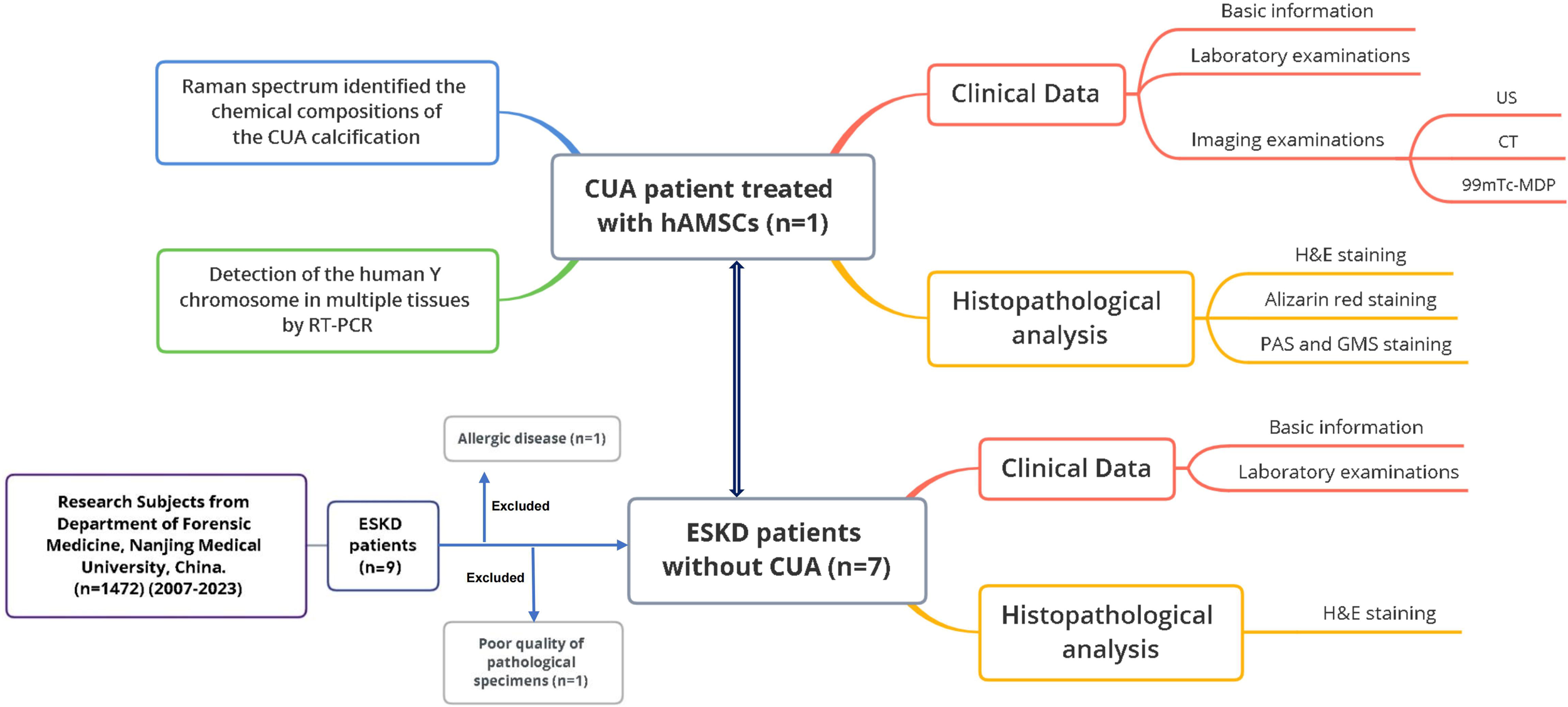
Flow diagram of the study. CUA, calcific uremic arteriolopathy; hAMSCs, human amnion–derived mesenchymal stem cells; ESKD, end-stage kidney disease; RT-PCR, reverse-transcription–polymerase chain reaction; US, ultrasound; CT, computed tomography; ^99m^Tc-MDP, whole-body scan with ^99m^Tc-methylene diphosphonate; H&E, hematoxylin and eosin staining; PAS, periodic acid-Schiff staining; and GMS, Grocott’s methenamine silver staining.

### Data analysis

Statistical analysis was performed using the IBM SPSS Statistics (version 22.0). Continuous variables are shown as mean values. Categorical variables are shown as frequencies and percentages.

## Results

### Study population

#### The CUA patient passed away due to a post-stroke intracerebral hemorrhage after 20 months of hAMSC treatment

The CUA patient had extensive necrotic ulcers with black eschars and infection on the lower extremities before hAMSC treatment (Figure 2A). The skin gradually healed after hAMSC treatment for 12 months (Figure 2B)^[18]^. The CUA patient passed away due to a post-stroke intracerebral hemorrhage after treatment with hAMSCs for 20 months. The integrity of the skin was still preserved (Figure 2C), and she had no obvious structural abnormalities in the skin tissues (Figure 2D–E)^[18]^.

**Figure 2:**
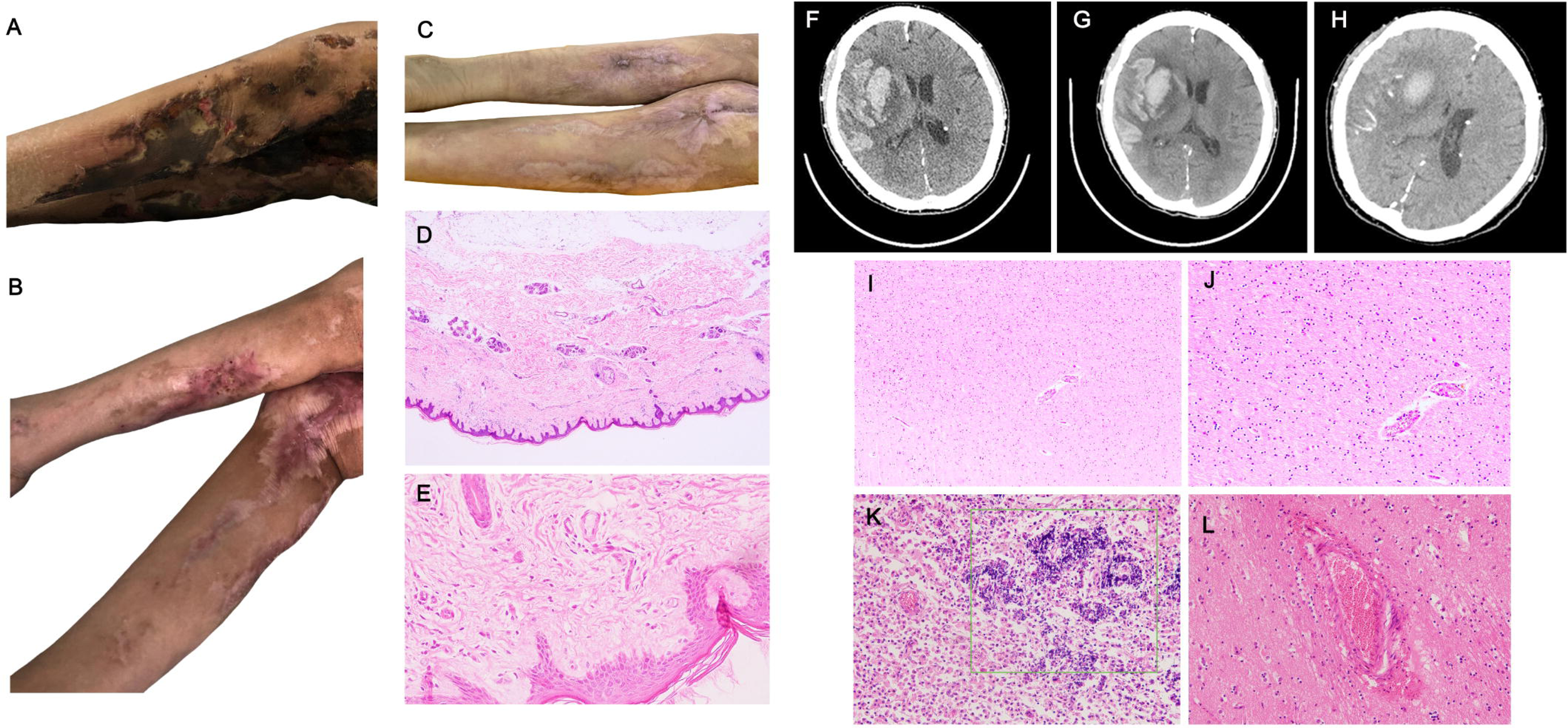
Skin tissues, plain head CT scans, and the brain tissues of the calciphylaxis patient treated with hAMSCs. (A–E) Skin tissues of the calciphylaxis patient. (A) Before hAMSC treatment, there were extensive necrotic ulcers with black eschars and infection on the lower extremities. (B) Most of the lower-limb skin tissues underwent regeneration, accompanied by local scar formation after hAMSC treatment for 12 months. (C) The skin ulcer healed, accompanied by the formation of local scars after hAMSC treatment for 20 months. (D–E) Twenty months after hAMSC treatment, the integrity of the skin structure has been preserved. The epidermal cells were arranged in well-organized multiple layers, while the dermis exhibited abundant collagen fibers. Moreover, the fiber bundles within the dermis displayed a staggered and woven pattern. There was an absence of inflammatory cells, microvascular calcification, and microthrombosis, indicating a positive outcome of the hAMSC treatment (D, H&E, 40×; E, H&E, 400×). (F–H) Plain head CT scans of the calciphylaxis patient. (F) May 2020: There was a cerebral gyrus hemorrhage in the right frontotemporal lobe, along with a patchy cerebral hemorrhage measuring 4.2 cm × 2.0 cm in the right basal ganglia. Edema was present in the surrounding brain tissue, and there were compression of the right lateral ventricle and a left deviation of the midline structure. (G) The next day: The edema of the surrounding brain tissue was more apparent. (H) After 12 days: The hemorrhage in the right frontotemporal lobe and patchy intracerebral areas was absorbed better than before, with the largest layer measuring 3 cm × 2.2 cm, resulting in reduced density. Additionally, the area of edema in the peripheral brain tissue was smaller than before. (I–L) Brain (H&E: I–J, the ESKD patient; K–L, the calciphylaxis patient). (I) There was no obvious structural abnormality (100×). (K) There were extensive inflammatory cells, including macrophages and plasma cells, and scattered calcification in the necrotic and encephalomalacia foci (100×). (J and L) The structures of the vessels appeared normal (200×). CT, computed tomography; hAMSCs, human amnion–derived mesenchymal stem cells; H&E, hematoxylin and eosin staining; and ESKD, end-stage kidney disease.

At the beginning of May 2020, the patient received the diagnosis of a fresh cerebral infarction at the local hospital but was not given thrombolysis or interventional therapy. In May 2020, the patient presented to our hospital, and a plain head computed tomography (CT) scan (Figure 2F) showed a hemorrhage in a cerebral gyrus of the right frontotemporal lobe and patchy cerebral hemorrhage in the right basal ganglia. The head CT scan in May 2020, revealed patchy intracerebral hemorrhage in the right basal ganglia (Figure 2H), with the largest layer measuring 3 cm × 2.2 cm, which had reduced density compared with that before (Figure 2F–G). Four days later, the patient suddenly showed signs of clenched teeth, binocular gaze, and opisthotonos. Though rescue measures were promptly taken, the patient passed away.

Supplementary Tables S1–S4 summarize the physical examination, laboratory examination, ultrasound examination, and computed tomography (CT) of the CUA patient before hAMSC treatment (July 2018) and after hAMSC treatment for 20 months (May 2020).

#### The ESKD patients

Table 1 presents the clinical data of ESKD patients. All of them were men, and their median age at death was 54.3 years. The causes of death in the ESKD patients included traffic accidents (n = 2), hyperkalemia (n = 2), head trauma (n = 1), heart failure (n = 1), and complications of thoracentesis (n = 1). Relevant comorbidities were hypertension (n = 5) and diabetes (n = 1).

**Table 1:**
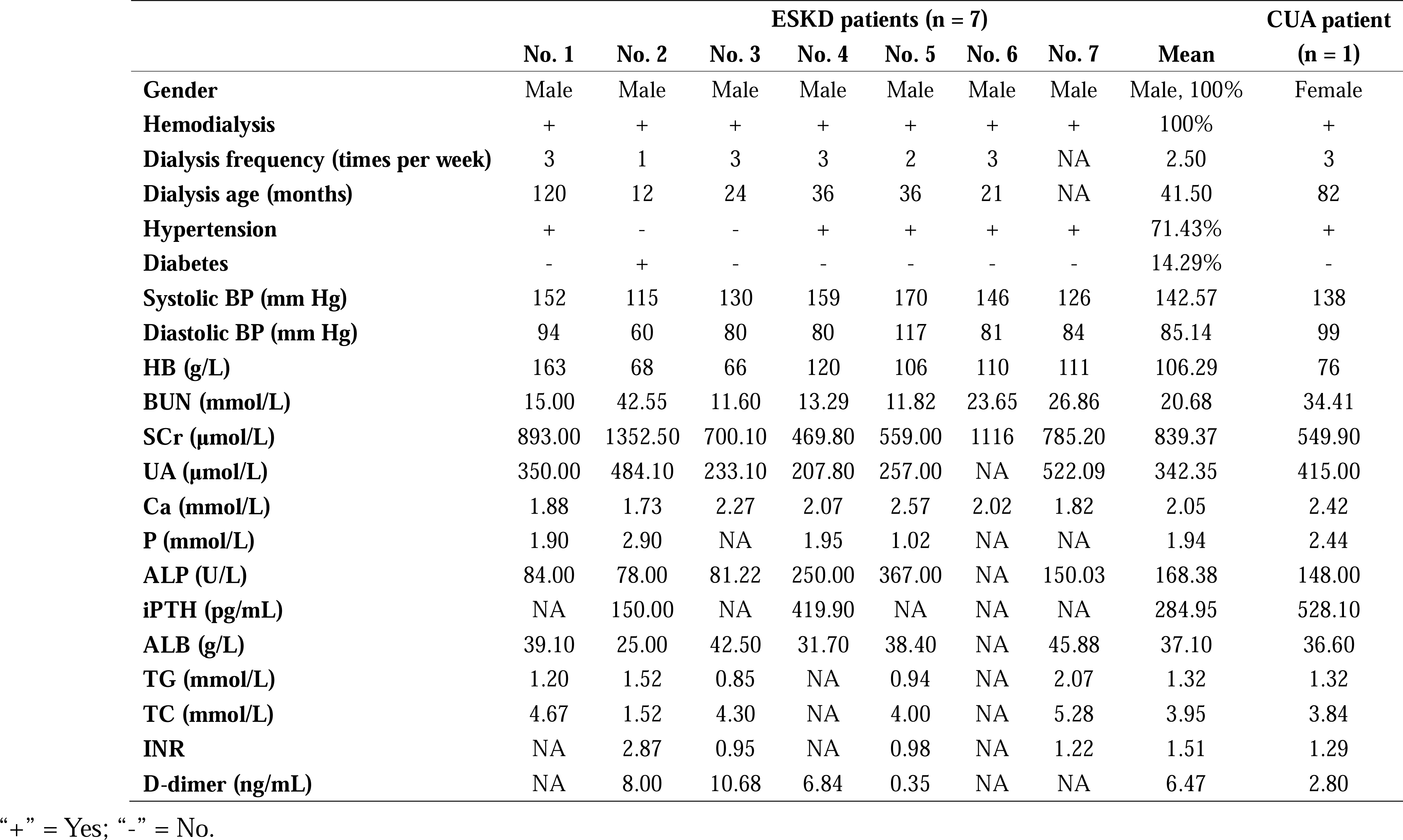
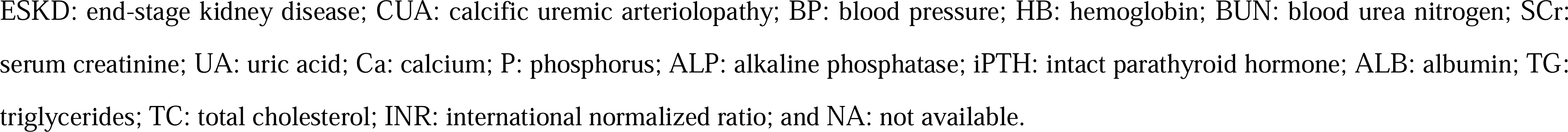
Demographic, clinical, and biochemical data of the CUA and ESKD patients.

### Comparisons of extracutaneous tissues between ESKD patients and the CUA patient treated with hAMSCs

#### Brain

At autopsy of the CUA patient, there were massive hemorrhage and encephalomalacia in the parenchyma of the right frontal lobe and temporal lobe, along with cerebral edema and cerebellar tonsillar herniation, which were considered to be the cause of death. No obvious structural abnormality was found in the brain tissues of the ESKD patients (Figure 2I–J). In the CUA patient, there were extensive inflammatory cells, including macrophages and plasma cells, and scattered calcifications in the necrotic and encephalomalacia foci (Figure 2K). The structures of vessels in the nearby hemorrhage area appeared normal (Figure 2L).

#### Cardiovascular system

Coronary atherosclerosis (7/7, 100%) and calcium deposition within the atheromatous plaque (4/7, 57%) were found in the ESKD patients (Figure 3A–B; Table 2). There were diffuse calcification and inflammatory cell infiltration of coronary arteries (Figure 3C–F) in the CUA patient treated with hAMSCs. Local thickening of the aortic wall and subintimal atherosclerotic plaque formation were noticed in the CUA patient (Figure 3G–H). The ESKD patients showed no obvious calcification or fibrosis in heart tissues (Figure 3I–J). In contrast, the CUA patient displayed marked fibrous hyperplasia and suspected scattered calcification loci within the myocardium (Figure 3K–M). Echocardiography revealed that there was a mass in the posterior lobe of the mitral valve in the CUA patient (Supplementary Video S1, please contact the corresponding author to request access to this video). In Figure 3N, the circle indicates the calcification of the posterior annulus of the mitral valve at that location, with mitral valve mucus calcification and degeneration (Figure 3O–P). The CUA patient had no inflammatory cell infiltration, bacteria, fungi, granulomas, or pneumocystis in the mitral valve (Figure 3Q–S).

**Figure 3:**
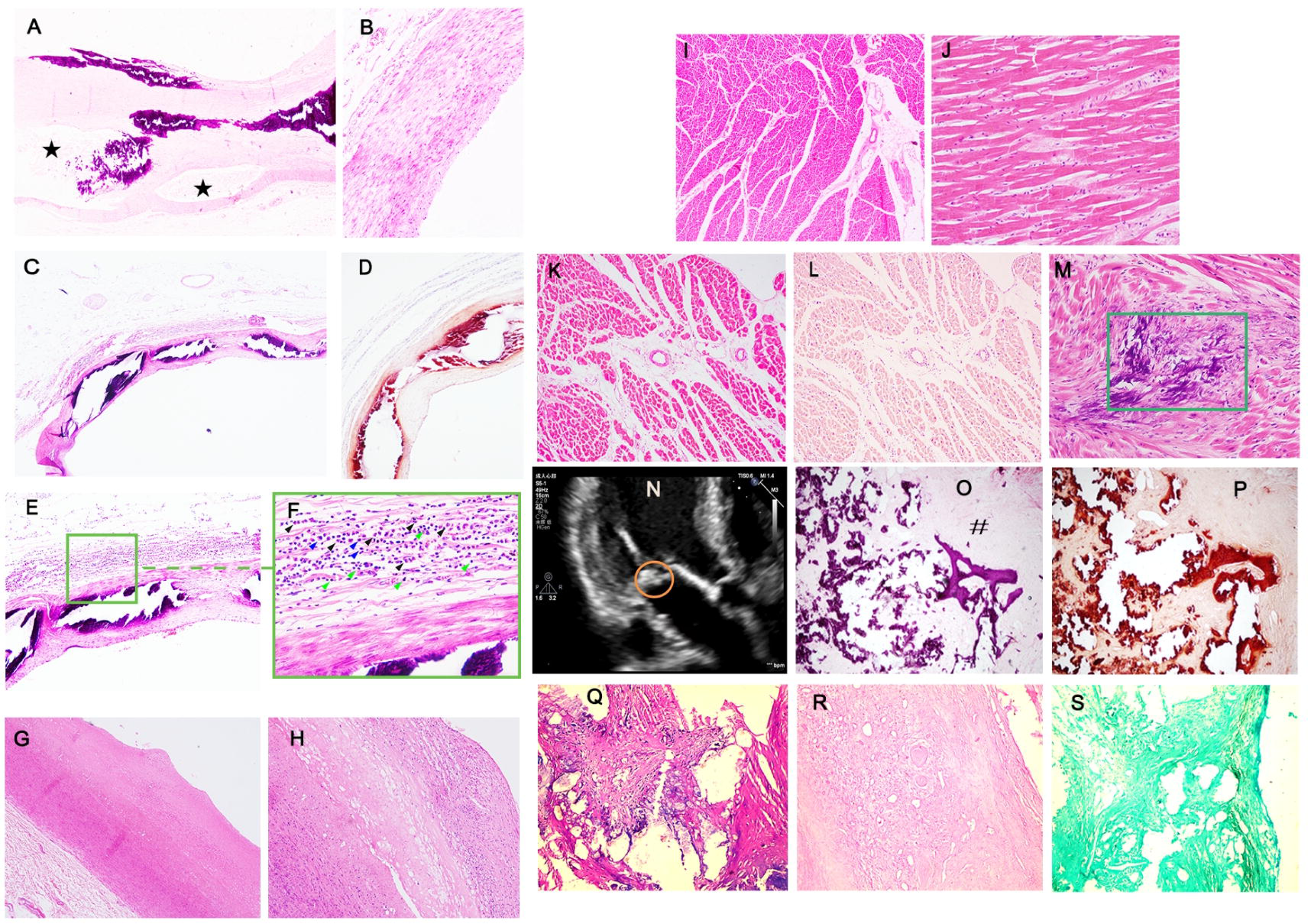
Comparison of the cardiovascular system from the ESKD patients and the calciphylaxis patient treated with hAMSCs. (A–B) ESKD patients (H&E). (A) Coronary atherosclerosis was characterized by unevenly thickened arterial walls, fibrous hyperplasia, calcium deposition within the atheromatous plaque, and formation of small vascular lacunae (*) (40×). (B) Thickened arterial wall and lack of inflammatory cell infiltration in the adventitia of the coronary artery wall (200×). (C–F) The calciphylaxis patient. (C) Diffuse medial circumferential calcification, fibrous intimal hyperplasia, and unevenly thickened arterial wall (H&E, 40×). (D) Medial circumferential calcification of the coronary artery wall (Alizarin red, 40×). (E) Diffuse inflammatory cell infiltration (H&E, 200×). (F) Lymphocytes (Δ), macrophages (Δ), and eosinophils (Δ) in the adventitia of the coronary artery wall (H&E, 400×). (G–H) Aorta tissues of the calciphylaxis patient (H&E). Local thickening of the aortic wall (G, 40×), subintimal atherosclerotic plaque formation, and inflammatory cell infiltration (H, 200×). (I–J) The ESKD patients (H&E). Normal histological structure (I, 40×) and slight fibrous hyperplasia within cardiomyocytes (J, 200×). (K–M) The calciphylaxis patient. (K) Marked fibrous hyperplasia within the myocardium (H&E, 40×). (L) No obvious calcification within the myocardium (Alizarin red, 40×). (M) Suspected scattered calcification loci and fibrous hyperplasia within the myocardium (H&E, 200×). (N–Q) The calciphylaxis patient. (N) Representative images of cardiac valve calcification by echocardiography, with the circle indicating the calcification of the posterior annulus of the mitral valve. (O) Mitral annular mucus degeneration (#), fibrous hyperplasia, calcification, and destroyed valve (H&E, 40×). (P) Mitral annular calcification (Alizarin red, 40×). (Q–S) Mitral valve (40×). No inflammatory cell infiltration (Q, H&E). No obvious colonies and hyphae (Q, H&E R, Periodic acid-Schiff). No bacteria, fungi, granulomas, or pneumocystis (S, Grocott’s methenamine silver). ESKD, end-stage kidney disease; hAMSCs, human amnion–derived mesenchymal stem cells; and H&E, hematoxylin and eosin staining.

**Table 2:**
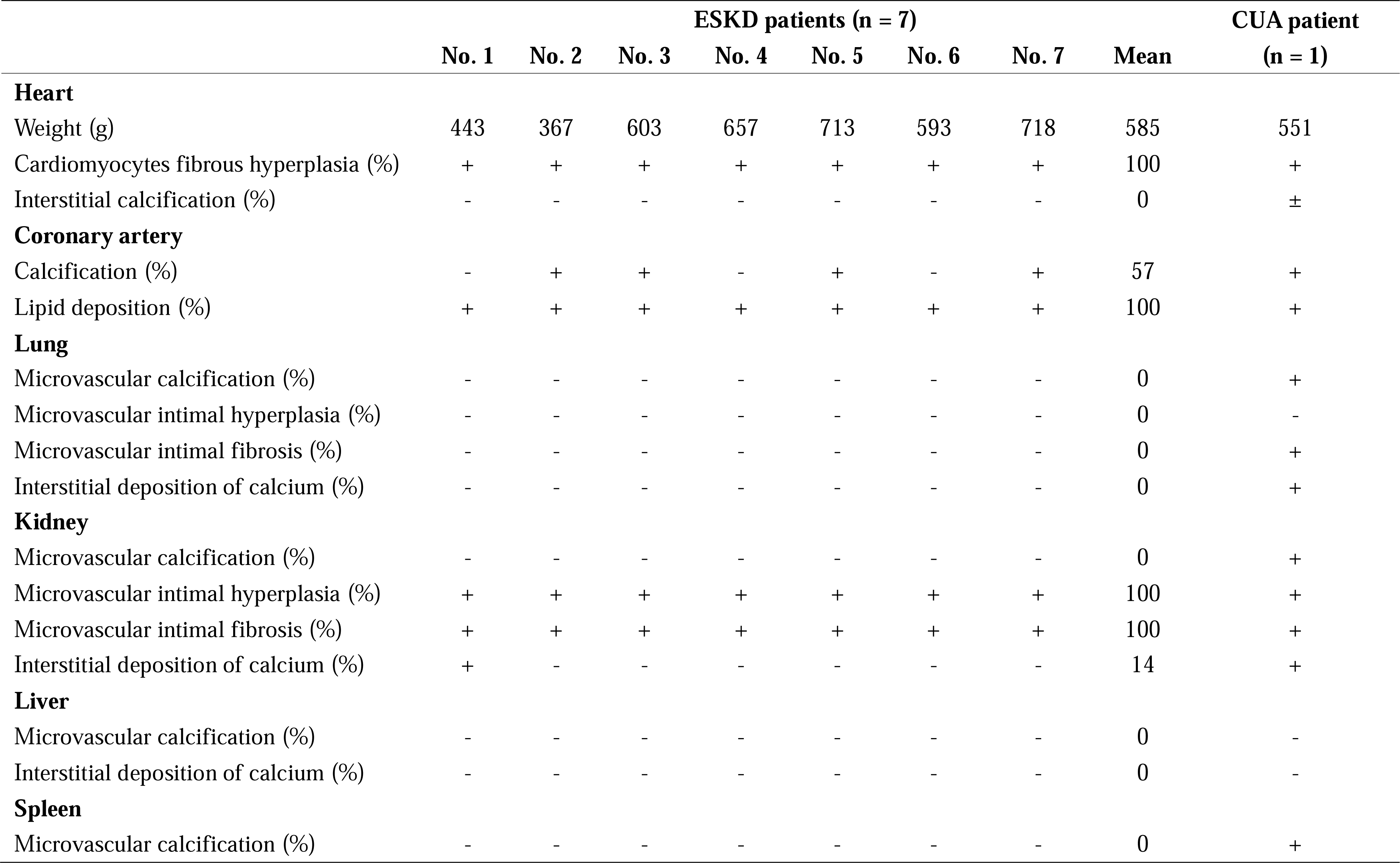
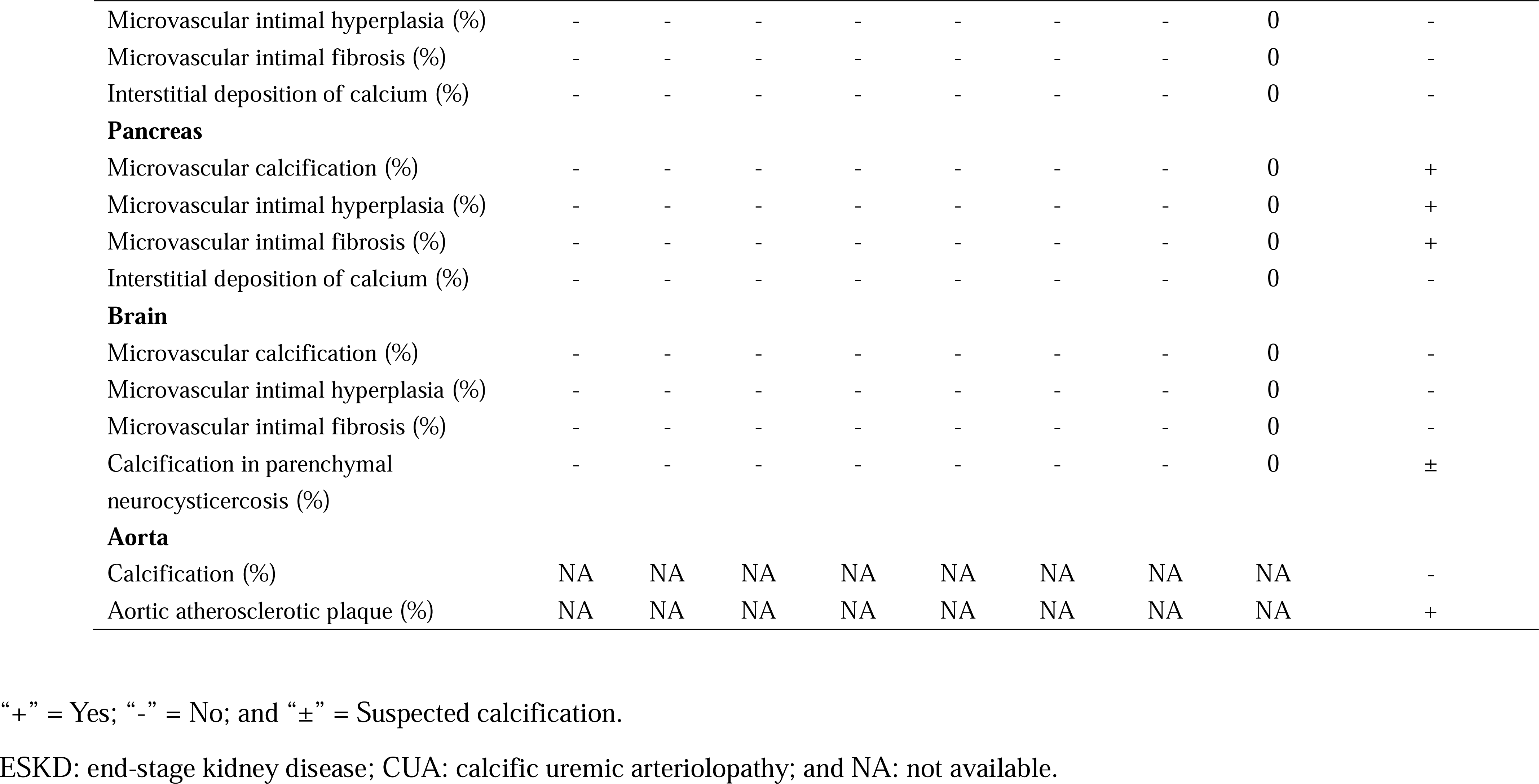
Histopathological features of multiple organs in the ESKD and CUA patients.

#### Lungs and kidneys

##### Whole-body bone scan

On admission, a whole-body bone scan with technetium-99m methylene diphosphonate (^99m^Tc-MDP) showed diffuse calcification in both the lungs and kidneys, as well as increased patchy uptake in the soft tissues of the thighs and legs, suggesting calciphylaxis (Figure 4A). Four months after hAMSC treatment, the whole-body bone scan showed a significantly decreased radiotracer uptake in the thighs and legs, indicating an improvement in skin lesions. In contrast, the uptake in the lungs and kidneys remained the same (Figure 4B). There were no typical findings of osteodystrophy due to secondary hyperparathyroidism on either early or later bone scan images (Figure 4A–B).

**Figure 4:**
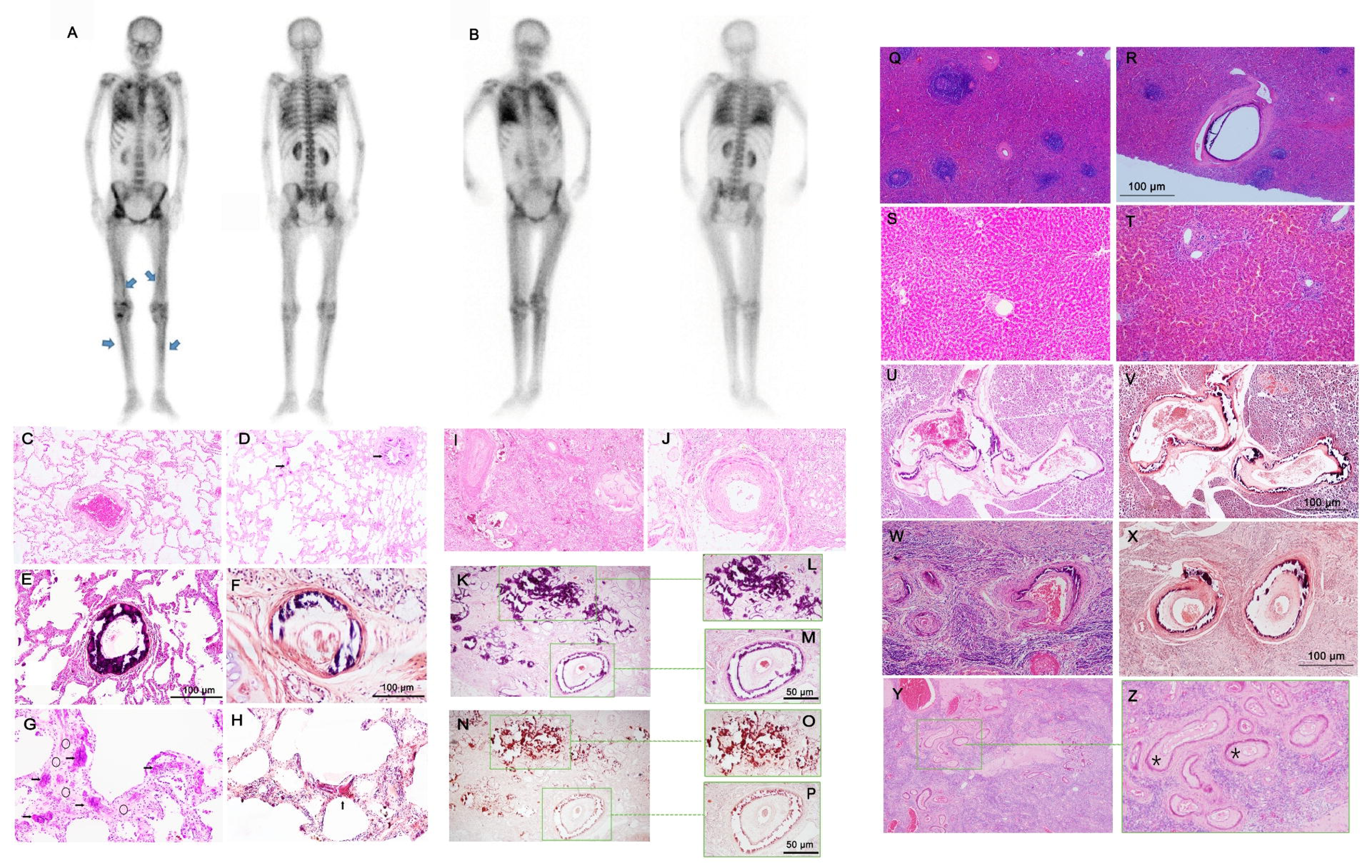
Histopathological analysis of multiple organs from the calciphylaxis patient treated with hAMSCs and the ESKD patients. (A–B) Whole-body bone scan with ^99m^Tc-methylene diphosphonate of the calciphylaxis patient. (A) Before hAMSC treatment, the radiotracer uptake was obvious in the lungs, kidneys, pelvis, medial thigh, and lateral calf. (B) Four months after hAMSC treatment, bone scintigraphy revealed minimal uptake in the medial thigh and lateral calf, but no obvious changes in the radiotracer uptake in the lungs, kidneys, and pelvis. (C) No obvious structural abnormality in the lung tissues of the ESKD patients (H&E, 40×). (D–H) The lung tissues of the calciphylaxis patient. (D) No obvious structural abnormality in the lung tissues, and slight calcific deposits in the bronchioles and alveolar septa (→) (Hɮ, 40×). (E–F) Microvascular medial circumferential calcification, intimal thickening, and destroyed vessel wall (E, Hɮ, 100×; F, Alizarin red, 400×). (G) Thickened alveolar septa with coarse calcification (→) and fibrosis (→) (Hɮ, 200×). (H) Calcific deposits in alveolar septa (→) Alizarin red, 200×). (I–J) The ESKD patients (H&E). (I) Extensive glomerular atrophy and interstitial fibrosis (40×). (J) Microvascular intimal thickening, fibrous hyperplasia, and lumen stenosis without calcification (200×). (K–P) The calciphylaxis patient (K–M, H&E N–P, Alizarin red). Microvascular intimal thickening, obvious lumen stenosis, medial circumferential calcification, and destroyed vessel wall (K, 40×; M, 200×). Calcification of glomeruli and renal tubules (L, 200×). (N–P) Diffuse calcium deposition in microvessels, glomeruli, and renal tubules (N, 40×; O–P, 200×). (Q–R) Spleen (H&E, 40×) of the CUA patient. (Q) Congested red pulp with atrophic white pulp. (R) Microvascular medial circumferential calcification, and destroyed vessel wall. (S–T) Liver (H&E, 100×). (S) No obvious structural abnormalities were found in the livers of the patients with ESKD. (T) The structure of hepatic lobules appeared normal in the calciphylaxis patient, with congestion in the hepatic sinusoids and mild inflammatory cell infiltration in the portal area. (U–V) The pancreas of the CUA patient. (U) Microvascular intimal fibrous hyperplasia, lumen stenosis, medial circumferential calcification, and destroyed vessel wall (H&E, 40×). (V) Microvascular medial circumferential calcification (Alizarin red, 40×). (W–X) Uterine tissues of the CUA patient. (W) Microvascular intimal fibrous hyperplasia, luminal stenosis, medial circumferential calcification, and destroyed vessel wall (H&E, 40×). (X) Microvascular medial circumferential calcification (Alizarin red, 40×). (Y–Z) Ovary tissues (H&E) of the CUA patient. Intimal thickening *), lumen stenosis, and lack of obvious calcification of microvessels (Y, 40×; Z, 100×). ESKD, end-stage kidney disease; hAMSCs, human amnion–derived mesenchymal stem cells; and H&E, hematoxylin and eosin staining.

##### Histological features

The ESKD patients showed no obvious structural abnormalities or calcific deposits in the microvessels and mesenchyme of lung and kidney tissues (Figure 4C, I–J). The CUA patient showed slight calcification of microvascular media and thickened alveolar septa with coarse calcification and fibrosis in lung tissues (Figure 4D–H).

In the ESKD patients, there was extensive glomerular atrophy and microvascular intimal thickening in the kidneys (Figure 4I–J). Notably, microvascular intimal thickening, obvious lumen stenosis, medial circumferential calcification, and diffuse calcium deposition in glomeruli and tubules were noted in the kidneys (Figure 4K–P).

### Spleen, liver, pancreas, and uterus

The ESKD patients showed no obvious structural abnormalities in the microvessels and mesenchyme of the spleen, liver (Figure 4S), and pancreas (Table 2). In the CUA patient, microvessel medial circumferential calcification and destroyed vessel wall were observed in the spleen, pancreas, and uterus (Figure 4R, U–X).

## Summary of the histopathological features of the extracutaneous tissues of ESKD and CUA patients

Table 2 lists the histopathological changes in the extracutaneous tissues of the CUA patient and those of the ESKD cases. Compared with ESKD patients, the CUA patient showed more significant lesions in the microvessels, such as calcification, hyperplasia, and fibrosis. However, we did not find any calcification of microvessels in the ESKD patients. The CUA patient showed interstitial calcification of the lungs and kidneys, particularly diffuse calcium deposition in glomeruli and tubules. However, only one ESKD patient with polycystic kidney disease exhibited interstitial calcification. Calcification of coronary arteries was evident in both groups.

The CUA patient demonstrated more obvious hyperplasia of microvessels in multiple extracutaneous tissues, including the kidneys, pancreas, uterus, and ovary, which was only evident in the kidneys of the ESKD patients. There was distinct intimal fibrosis of microvessels in multiple extracutaneous tissues of the CUA patient, including the lungs, kidneys, pancreas, uterus, and ovary. In contrast, the ESKD patients only had intimal fibrosis of the microvessels in the kidneys.

Although intra- and extravascular calcium deposition was noted in multiple extracutaneous tissues, associated tissue micro-thrombus was not found in the CUA patient. Furthermore, no tumor or abnormal hyperplasia was found in multiple tissues of the CUA patient, including the brain, hypophysis, parathyroid gland, heart, cardiac valve, coronary artery, aorta, lungs, liver, spleen, pancreas, kidneys, uterus, ovary, adipose tissues, and skin tissues.

## Raman spectrum analysis

Raman spectra of calcifications of the mitral valve and kidneys of the CUA patient were collected. By comparing the Raman spectra with previous studies^[26,27]^, it was revealed that calcifications of the mitral valve and kidney tissues were composed of calcium phosphate and a small amount of calcium carbonate (Figure 5A–C). The morphological feature of calcification was lumpy, with the highest density in the middle and gradually decreasing around the circumference (Figure 5D–E).

**Figure 5:**
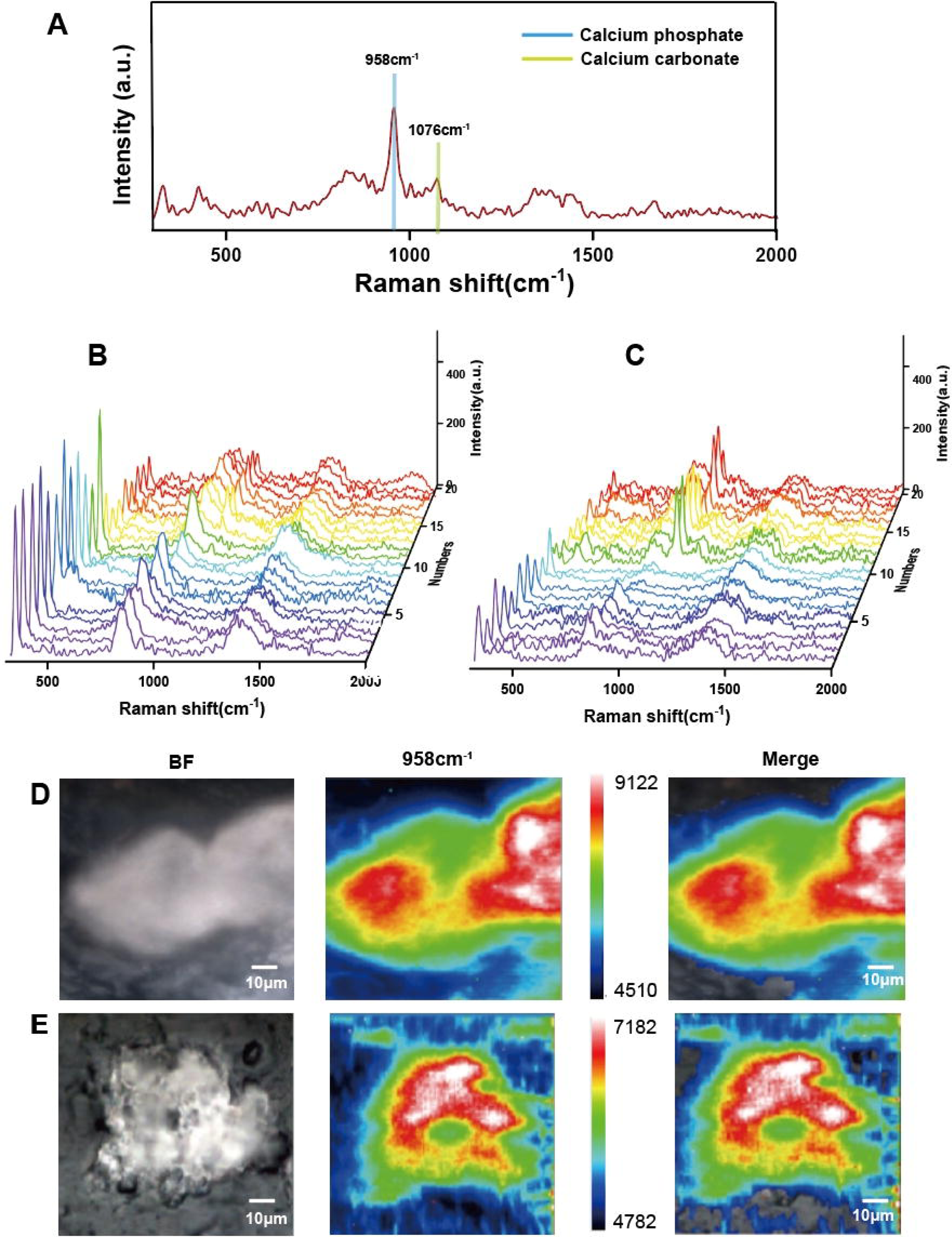
Raman mapping of calcium deposition in the calciphylaxis patient treated with hAMSCs. (A) Raman spectral data corresponding to calcium phosphate (958 cm^−1^) and calcium carbonate (1076 cm^−1^) in the mitral valve and kidney tissue. (B) Raman spectra of 20 random calcium depositions in the mitral valve tissues. (C) Raman spectra of 20 random calcium depositions in the kidney tissues. (D) Raman mapping of calcium phosphate in the mitral valve (1000×). (E) Raman mapping of calcium phosphate in the kidney tissue (1000×). hAMSCs, human amnion–derived mesenchymal stem cells; BF, bright field.

## Expression of the human Y chromosome in the extracutaneous tissues of the CUA patient

The expression of the Y chromosome was not detected by RT-PCR in brain tissues, hypophysis, parathyroid gland, spleen, lungs, cardiac valve, coronary artery, ovary, adipose tissues, skin of the thigh, and dermal tissue of the buttocks in the CUA patient after treatment with hAMSCs for 20 months (Supplementary Figure S1).

## Discussion

In 1961, Seyle first proposed the concept of “calciphylaxis,” which described the presence of calcium deposition in skin tissue of uremic rats after stimulation by sensitizing agents, without observing skin ischemic necrosis^[28]^. However, calciphylaxis in humans is not an inducible hypersensitivity reaction disease. Currently, no animal model for calciphylaxis corresponds to clinical presentation^[29,30]^. According to an assessment of 56 ESKD patients by autopsy, Kuzela et al. found that the incidence of severe calcification of the heart, lungs, stomach, and kidneys was 33.93%, 48.21%, 21.43%, and 30.19%, respectively^[31]^. In our study, calcification of the coronary arteries was observed in both groups. There was no prevalent calcification of microvessels in the extracutaneous tissues of the ESKD patients. The incidence of calcification of the kidneys of our ESKD group was 14.29%, which is lower than that reported in the literature (30.19%^[31]^ and 67%^[32]^).

According to autopsy reports of three calciphylaxis patients^[33]^, all individuals had cardiovascular calcification. Vascular calcification of the pancreas and kidneys was noted, but there was no indication of calciphylaxis in other tissues. The authors concluded that calciphylaxis exclusively affects the skin and does not involve extracutaneous tissues^[33]^. A study of 36 CUA patients showed that the calcification was always located circumferentially, mostly in the intima and media of the skin vessels, and often associated with interstitial deposits^[26]^. Medial circumferential calcification was found in the skin, thymus, and kidney vessels in calciphylaxis patients^[9,33,34]^. There is mounting evidence that calciphylaxis affects extracutaneous tissues, such as the lungs, kidneys, heart, and mesentery^[8,10,13,35–37]^.

In our research, diffuse medial calcification of extracutaneous tissues, including the lungs, kidneys, spleen, pancreas, and uterus, was more prevalent in the CUA patient treated with hAMSCs than in the ESKD patients. The CUA patient underwent hemodialysis with the utilization of a long-term catheter, specifically inserted into the right internal jugular vein. This procedure posed an increased risk of heart valve infection. However, echocardiography imaging and histological special staining indicated a significant level of calcification on the posterior annulus of the mitral valve. These findings ruled out the occurrence of acute cerebral infarction in May 2020, which was previously hypothesized to be caused by the embolism resulting from the detachment of valvular bacterial thrombus.

Determining the chemical composition of calcifications involving extracutaneous tissues could shed light on the pathogenesis of calciphylaxis. A clear and straightforward boundary of calcifications can be obtained through Raman mapping^[25]^. Colboc et al. found that the skin calcification in calciphylaxis patients was composed of pure calcium-phosphate apatite, as determined by Raman spectroscopy^[26]^. Lloyd et al. confirmed the presence of carbonated apatite in calciphylaxis tissues using Raman spectroscopy and microcomputed tomography^[27]^. We revealed that extracutaneous calcifications of the mitral valve and kidney tissues of the CUA patient were mainly composed of calcium phosphate and a small amount of calcium carbonate.

From an external perspective, hAMSC treatment had the ability to enhance the healing process of the skin and soft tissues in this CUA patient. Regarding the lung tissues, there were no obvious changes in increased lung uptake after treatment with hAMSCs for 4 months, as shown by whole-body bone scintigraphy. This indicates obvious microvessel calcification in lung tissues. However, after 20 months of hAMSC treatment, histopathological analysis of the lung tissues in the CUA patient revealed only slight calcific deposits in the bronchioles and alveolar septa. Of note, there was still obvious calcification in other extracutaneous tissues, including the mitral valve, kidneys, spleen, pancreas, and uterus. Recent data indicate that a significant portion of administered stem cells become initially trapped in the lungs, a phenomenon known as the first-pass effect^[38]^. This observation is supposed to have a connection with the diverse extracutaneous histopathological findings seen in the CUA patient who underwent hAMSC treatment.

It has been reported that 60% of calciphylaxis patients have severe thrombophilia^[39]^. CUA patients (n = 34) are significantly more likely to show thrombi (82%), including focal (32%) and diffuse (50%) thrombi, than ESKD patients (n = 26)^[40]^. Thrombosis is one of the key factors in the diagnosis of calciphylaxis, which is associated with a pro-thrombotic local environment that predisposes to vascular occlusion^[1,41]^. We demonstrated that the CUA patient exhibited signs of hypercoagulability prior to the administration of hAMSCs, including elevated platelet aggregation levels, D-dimer, and fibrinogen^[17]^. After the administration of hAMSCs, significant improvements were observed in coagulation markers in this particular case^[17]^. It is worth noting that no thrombosis was detected in the extracutaneous tissues of the CUA patient who underwent hAMSC treatment, further supporting the notion that hAMSCs can ameliorate the hypercoagulability associated with CUA^[17]^.

Following intravenous injection, hAMSCs promptly exit the bloodstream and establish colonies in various tissues and organs^[42]^. Our animal experiments demonstrated that hAMSC DNA was detected in the lungs, kidneys, and muscle tissues of the mice 1 month after receiving hAMSC injection^[17]^. This finding suggests that hAMSCs can migrate to and reside in multiple tissues^[17]^. However, over time, these cells undergo apoptosis^[43]^. In this study, by detecting the expression of the Y chromosome, we did not find the transplanted hAMSCs in multiple extracutaneous tissues of the CUA patient after 20 months of stem cell treatment.

Han et al. reported a distinctive case of immature teratoma in a patient who underwent human iPSC-derived cell therapy for diabetes^[19]^. It is important to note that hAMSCs are non-tumorigenic^[44]^, and one of the reasons for their safety profile is the absence of cell surface markers associated with tumorigenesis, such as CD34, CD133, and HLA-DR^[42]^. Moreover, hAMSCs express minimal levels of HLA-A, -B, and -C^[42,45]^. Immunosuppressants are not required during hAMSC treatment.

We observed that the tumor biomarkers of this CUA patient remained within normal ranges during the entire follow-up period^[18]^. Moreover, we found no evidence of tumor cell presence or abnormal tissue hyperplasia in the various extracutaneous tissues analyzed by histology in the hAMSC-treated CUA patient. These findings indicate that the prolonged and frequent administration of hAMSCs to the patient did not increase the likelihood of tumor formation. However, in light of the potential risks involved, it is advisable to closely monitor tumor biomarkers in patients undergoing hAMSC treatment.

There are several limitations to this study. First, calciphylaxis is a rare disease, which led to the inclusion of only one CUA patient who received treatment with hAMSCs and subsequently underwent autopsy. Second, this study did not incorporate a comprehensive pathological analysis of multiple tissues prior to the initiation of hAMSC treatment. This limitation is attributed to the impracticality of conducting a pathological analysis on all the patient’s tissues prior to commencing therapy. Third, this study did not include whole-body bone scintigraphy at the end of the follow-up period as the patient’s sudden cerebral hemorrhage resulted in her unfortunate demise. In conclusion, while hAMSC therapy was proven beneficial in terms of wound healing in this CUA patient, it did not completely alleviate the systemic pathology associated with calciphylaxis, especially vascular injury. The microvascular calcifications were composed of calcium phosphate and calcium carbonate. No tumorigenicity was observed. Further research is necessary to comprehend the underlying mechanisms and develop more efficient individualized hAMSC treatment strategies that are guided by biomarkers. This research not only concentrates on the potential applications of hAMSC treatment for skin lesions but also investigates its efficacy in protecting internal organs.

## Data Availability

All data produced in the present work are contained in the manuscript

https://www.example.com

## Availability of data and materials

The data that support the findings of this study are available from the corresponding author upon reasonable request.

## Supplementary material

**Supplementary File (PDF)**

**Supplementary Table S1:** Comparison of physical examinations of the CUA patient before and after hAMSC treatment.

**Supplementary Table S2:** Comparison of laboratory parameters of the CUA patient before and after hAMSC treatment.

**Supplementary Table S3: Comparison of ultrasound examinations of the CUA patient before and after hAMSC treatment.**

**Supplementary Table S4:** Comparison of CT examinations of the CUA patient before and after hAMSC treatment.

**Supplementary Figure S1:** Twenty months after hAMSC treatment, the presence of the Y chromosome was detected in multiple tissues of the female calciphylaxis patient through RT-PCR.

