## Supplementary File for "Histopathology of extracutaneous tissues in a uremic calciphylaxis patient treated with human amnion-derived mesenchymal stem cells: a comparison with ESKD patients"

**SUPPLEMENTARY MATERIAL**

**Supplementary Material Table of Contents**

### Supplementary Table S1: Comparison of physical examinations of the CUA patient before and after hAMSC treatment.

|  | **Before hAMSC treatment (****July 2018)** | **After hAMSC treatment (May 2020)** |
| --- | --- | --- |
| **Temperature, heart rate, respiratory rate, and blood pressure** | 37.4°C, 110 bpm, 18 bpm, and 134/76 mm Hg | 36.0°C, 133 bpm, 42 bpm, and 104/83 mm Hg |
| **General** | Awake and responsive, with emaciation and a severe anemia appearance | Somnolence, poor nutrition, emaciation, and an anemic appearance, but the condition has improved relative to that in 2018 |
| **Heent** | Facial edema | No facial edema; other items were the same as those in July 2018 |
| **Neck** | Supple; the right internal jugular vein was temporarily catheterized with full internal jugular venous; the trachea was in the middle; and there was no enlarged thyroid gland | The right internal jugular vein was not temporarily catheterized; other items remained the same as those in July 2018 |
| **Heart** | The cardiac boundary was expanded to the left and down; the heart rate was 110 bpm with an irregular rhythm; and the auscultation areas for the tricuspid and mitral valves revealed a 4/6 systolic rough murmur, which was heard and transmitted to the sternum and armpit | The heart rate was 133 bpm with an irregular rhythm; the tricuspid and mitral valve auscultation areas could be heard with a 3/6 systolic rough murmur; and other items were the same as those in July 2018 |
| **Lungs** | Both lungs’ breathing sounds were rough, with moist rales coming from the right lung, but no rhonchus could be heard | Thick respiratory sounds were heard from both lungs, with no obvious dry or wet rales |
| **Abdomen** | Soft, with no tenderness or rebound pain in the entire abdomen; Murphy’s sign was negative; the liver and spleen were not palpable under the ribs, and shifting dullness was absent; and there was no knocking pain in the kidney area | Same as in July 2018 |
| **Extremities** | Patchy ecchymosis was scattered on the buttocks, back, and thighs of the lower extremities; some parts of the skin showed painful ulcers that have formed scabs with purulent secretions; and the skin on the thighs was black, locally hardened, and tender | The skin lesions had almost healed; skin scars were scattered on the buttocks, back, and thighs of the lower extremities |
| **Skin** | At the entrance of the PD tunnel, there were skin ulcers with purulent secretions measuring 3 cm in length; pressure sores were visible in the caudal sacrum, with ulcers and scabs developed in the vulva; and scab with purulent secretions had formed on the skin of the lower abdomen and upper back | The skin lesions had almost healed, except for small areas of sacrococcygeal ulcers; skin scars were visible on the limbs, lower abdomen, and upper back |
| **Neurological exam** | The tension of upper-limb muscles was normal, and the tension of lower-limb muscles was between strength V and normal; there was knee joint stiffness with limited extension; and reflexes were physiological, with no pathological reflexes | Grade 0 muscle strength was detected in the left upper and lower limbs, while grade 5 muscle strength was observed in the right upper and lower limbs; the patient did not cooperate during the assessment of ataxia and muscular tension; and physiological reflexes were present, and the Babinski sign was positive on the left side |

CUA: calcific uremic arteriolopathy; hAMSCs: human amnion–derived mesenchymal stem cells; Heent: head, eyes, ears, nose, and throat.

### Supplementary Table S2: Comparison of laboratory parameters of the CUA patient before and after hAMSC treatment.

|  | **Before hAMSCs treatment (July 2018)** | **After hAMSCs treatment**  **(May 2020)** |
| --- | --- | --- |
| **WBC** **(× 10^9^/L)** | 12.03 | 8.77 |
| **NE (%)** | 80.0 | 76.5 |
| **HB (g/L)** | 55 | 76 |
| **Plt (× 10^9^/L)** | 555 | 331 |
| **TP (g/L)** | 62.8 | 67.7 |
| **ALB (g/L)** | 28.5 | 36.6 |
| **Ca (mmol/L)** | 2.24 | 2.42 |
| **P (mmol/L)** | 1.18 | 2.44 |
| **ALP** **(U/L)** | 288.1 | 148 |
| **iPTH (pg/mL)** | 528.1 | 455.9 |
| **CRP (mg/L)** | >90 | 15.3 |
| **PCT (ng/mL)** | 11.57 | 4.13 |
| **ANA** | (-) | (-) |
| **BC** | (-) | (-) |
| **BNP (pg/mL)** | >35,000 | >35,000 |
| **CF** | Prothrombin time: 11.40 s;  partial thromboplastin activation time: 23.10 s;  fibrinogen: 3.22 g/L;  thrombin time: 14.20 s; and  D-dimer: 1.5 mg/L | Prothrombin time: 14.60 s;  partial thromboplastin activation time: 27.60 s;  fibrinogen: 4.10 g/L;  thrombin time: 17.10 s; and  D-dimer: 2.8 mg/L |

CUA: calcific uremic arteriolopathy; hAMSCs: human amnion–derived mesenchymal stem cells; WBC: white blood cells; NE: neutrophils; HB: hemoglobin; Plt: platelets; TP: total protein; ALB: albumin; Ca: calcium; P: phosphorus; ALP: alkaline phosphatase; iPTH: intact parathyroid hormone; CRP: C-reactive protein; PCT: procalcitonin; ANA: antinuclear antibody; BC: blood culture; BNP: B-type brain natriuretic peptide; and CF: coagulation function.

### Supplementary Table S3: Comparison of ultrasound examinations of the CUA patient before and after hAMSC treatment.

|  | **Before hAMSC treatment**  **(July 2018)** | **After hAMSC treatment**  **(May 2020)** |
| --- | --- | --- |
| **Heart** | The enlargement of the whole heart (LAD 43 mm, LVDd 55 mm, LVDs 43 mm, RAD 42 mm, and RVDd 45 mm) with cardiac insufficiency (EF 43.7%); mitral and tricuspid valve calcification with moderate to severe regurgitation; and mild hypertension of the pulmonary artery (40 mm Hg by CW estimation) | There was left-ventricular hypertrophy (LAD 25 mm, LVDd 50 mm, LVDs 40 mm, RAD 30 mm, and RVDd 39 mm) with cardiac insufficiency (EF 36.0%); there were mild tricuspid regurgitation and mild mitral insufficiency; there was a mass in the posterior lobe of the mitral valve; and the pulmonary arterial pressure was estimated to be 21 mm Hg by CW estimation |
| **Internal and external iliac arteries** | No obvious abnormalities | NA |
| **Internal and external iliac veins** | No obvious thrombosis in veins | NA |

CUA: calcific uremic arteriolopathy; hAMSCs: human amnion–derived mesenchymal stem cells; LAD: left-atrial diameter (mm); LVDd: left-ventricular diastolic diameter (mm); LVDs: left-ventricular systolic diameter (mm); RAD: right-atrial diameter (mm); RVDd: right-ventricular diastolic diameter (mm); EF: ejection fraction (%); CW: continuous wave; and NA: not available.

### Supplementary Table S4: Comparison of CT examinations of the CUA patient before and after hAMSC treatment.

|  | **Before hAMSC treatment (July 2018)** | **After hAMSC treatment (May 2020)** |
| --- | --- | --- |
| **Lungs** | Both lungs showed diffuse ground-glass nodules and patchy, blurred shadows, with a small amount of pleural effusion on both sides | Diffuse ground glass and nodular shadow were present in both lungs |
| **Heart** | Enlargement of the heart shadow | Enlarged heart shadow, high-density shadow in the mitral valve area |
| **Liver** | Normal aspect and size without focal lesions; normal intrahepatic ducts | Small, round, low-density shadow in the left lobe of the liver, approximately 5 mm in diameter; other items were the same as those in July 2018 |
| **Gall bladder** | Normal-sized but with a lightly thickened gallbladder wall | Same as in July 2018 |
| **Pancreas** | Aspect, size, and density were normal without focal lesions; the pancreatic duct was not dilated; and the fatty space around the pancreas was clear | Same as in July 2018 |
| **Spleen** | Normal aspect, size, and density without focal lesions | Same as in July 2018 |
| **Kidney** | Bilateral atrophied kidneys with multiple low-density shadows, without obvious signs of enlargement or stasis in the ureters, renal pelvises, and calyxes; the perirenal fatty space was clear | Same as in July 2018 |
| **Vessels** | Multiple collateral circulations (calcified small blood vessels cannot be ruled out) were formed in the chest, abdominal wall, and abdominal soft tissue | Same as in July 2018 |

CUA: calcific uremic arteriolopathy; hAMSCs: human amnion–derived mesenchymal stem cells.

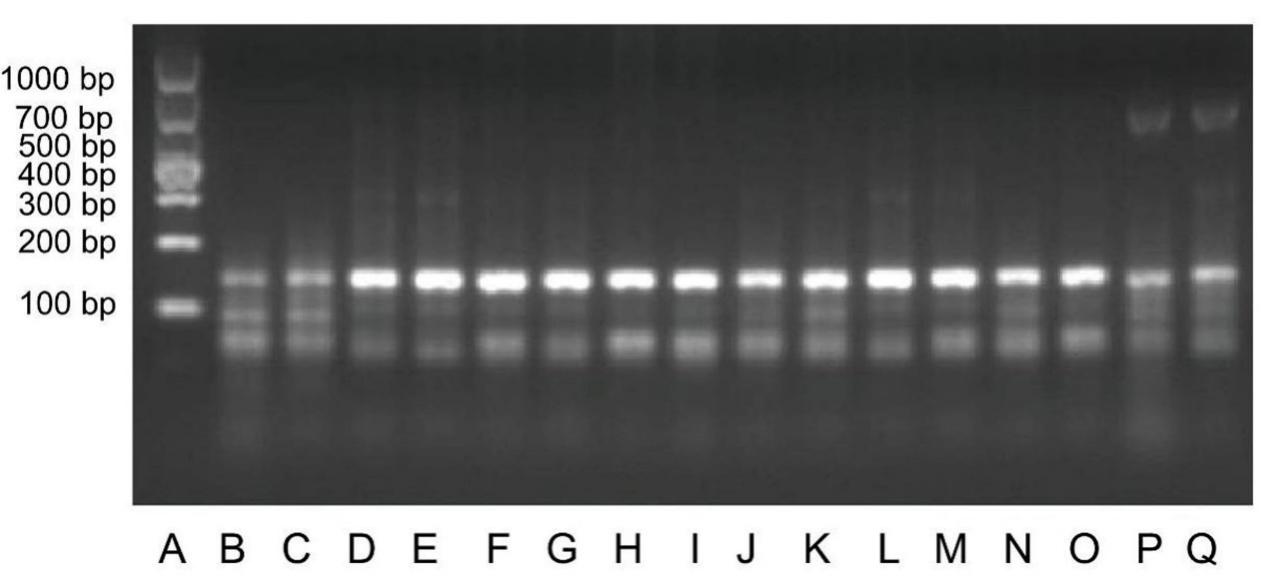

### Supplementary Figure S1: Twenty months after hAMSC treatment, the presence of the Y chromosome was detected in multiple tissues of the female calciphylaxis patient through RT-PCR.

(A) Marker-DL 1000; (B–C) normal female controls; (D) brain tissue surrounding an old cerebral hemorrhage; (E) brain tissue around a fresh cerebral hemorrhage; (F) [hypophysis](javascript:%20void(0)); (G) parathyroid; (H) spleen; (I) lung; (J) cardiac valves; (K) coronary artery; (L) ovaries; (M) adipose tissue; (N) skin of the thigh; (O) dermal tissue of buttocks; and (P–Q) normal male controls.

hAMSCs: human amnion–derived mesenchymal stem cells; RT-PCR: reverse-transcription–polymerase chain reaction.

### Supplementary Video S1: Echocardiography of the calciphylaxis patient treated with hAMSCs.

A calcified mobile mass in the posterior annulus of the mitral valve.

hAMSCs: human amnion–derived mesenchymal stem cells.
